# Respiratory syncytial virus-associated hospital admissions by deprivation levels among children and adults in Scotland

**DOI:** 10.1101/2023.08.15.23294119

**Authors:** Richard Osei-Yeboah, Fuyu Zhu, Xin Wang, Harish Nair, Harry Campbell, PROMISE investigators

## Abstract

**Background:** Socioeconomic deprivation may predispose individuals to respiratory tract infections (RTI). We aimed to estimate the number and rate of RSV-associated hospitalisations by socioeconomic status using the Scottish Index of Multiple Deprivation (SIMD).

**Methods:** Using national routine healthcare records and virological surveillance from 2010-2016, we used a time-series linear regression model and a direct measurement based on ICD-10 coded diagnoses to estimate RSV-associated hospitalisations by SIMD level and age and compared to influenza-associated hospitalisations.

**Results:** Using the model-based approach, we estimated an annual average rate per 1000 of 0.76 (95%CI: 0.43-0.90) for individuals of all ages in the least deprived group (5^th^ quintile of SIMD) to 1.51 (1.03-1.79) for the most deprived group (1^st^ quintile). Compared with the least deprived group, we estimated that the rate ratio (RR) was 1.96 (95%CI: 1.23-3.25), 1.60 (1.0- 2.66), 1.35 (0.85-2.25), and 1.12 (0.7-1.85) in the 1^st^ to 4^th^ quintile. The pattern of RSV-associated RTI hospitalisation rates variation with SIMD was most pronounced in children aged 2 years and below. The ICD-10 direct measurement approach provided much lower rates than the model-based approach but yielded similar RR estimates between SIMD groups.

**Conclusion:** Higher RSV hospitalisation rates are seen in the Scottish population of higher deprived levels. The differences between deprivation levels are most pronounced in infants and young children.

## Background

Respiratory syncytial virus (RSV) is a common cause of respiratory infections, causing substantial hospitalisations and deaths, especially in young children and the elderly [1, 2]. We estimated that globally in 2019 there were 33 million episodes and 3.6 million hospitalisations of RSV-associated acute lower respiratory infections in children younger than 5 years [3]. We have previously reported that there are about 245,000 [4] and 160,000 [5] RSV-associated hospitalisations annually in children under 5 years old and adults above 18 years old respectively in the European Union (EU) plus Norway and the United Kingdom. About 75% of hospitalisations in children under 5 years occur in infants (aged below 1 year) [4], while about 92% of hospitalisations in adults occur in those above 65 years [5].

Studies suggest that socioeconomic status [6, 7] is one of the key risk factors for respiratory infections, and the higher risks are not only restricted to low-income countries but are also present in poor and disadvantaged populations within the middle- and high-income countries [8]. Lewis et al. identified variations in the seasonality of bronchiolitis hospitalisations by socioeconomic level in England such that increased deprivation was found to be associated with less seasonal variation and a slightly delayed epidemic peak [9]. A study conducted in England showed that the risk of bronchiolitis hospitalisation was 38% greater for infants of the most deprived socioeconomic group at peak admission week compared with the least deprived group [9]. Hungerford et al. found that among adults, hospitalisations for influenza-associated illnesses were more frequent in the most socioeconomically deprived areas compared with the least deprived areas in North-West of England whereas, the rates in children were more homogenous across the socioeconomic strata [10].

Understanding the burden of RSV-associated illnesses, especially severe illnesses by deprivation levels would be useful for recommendations, guidance, and decisions on RSV immunisation strategies. In this regard, we aimed to estimate the average annual number and rates of RSV-associated respiratory tract infection (RTI) hospitalisations and influenza-associated RTI hospitalisations in children and adults based on socioeconomic status using the Scottish Index of Multiple Deprivation (SIMD).

## Methods

### Study design and population

The study design and data source have been described previously [11]. Briefly, we conducted a retrospective analysis of RSV-associated and influenza-associated RTI hospitalisations using Scottish national hospital registries during six consecutive epidemiological years (2010-2016). An epidemiological year included the period from week 40 of one year to week 39 of the next year. The study population included individuals hospitalised with RTI and recorded in the Scottish Morbidity Record 01 (SMR01), a Scottish national healthcare registry.

### Case definitions

As done previously [11, 12], we defined the incidence of RTI hospitalisations based on International Classification of Diseases – 10^th^ edition (ICD-10) diagnosis codes (Supplementary Table 1). RTI hospitalisation was defined as a hospital episode with any mention of RTI in the diagnosis codes either as a main or secondary diagnosis. RSV-RTI admission was RTI admission with any mention of an RSV ICD-10 diagnosis code indicating RSV either as a main or secondary diagnosis (Supplementary Table 1). Influenza-associated RTI hospitalisation was RTI admission with any mention of influenza ICD-10 diagnosis code indicating influenza either as a main or secondary diagnosis (Supplementary Table 1). Virological surveillance data sources

The Electronic Communication of Surveillance in Scotland (ECOSS) system captures laboratory results from all diagnostic and reference laboratories in Scotland. All positive RSV and influenza test results are included, though there is no denominator information on the tested population. Reliable data on RSV-positive confirmations are available from 2009 onwards [12, 13].

### Scottish Index of Multiple Deprivation (SIMD)

The SIMD is the Scottish Government’s tool for identifying the concentration of deprivation across Scotland. It is derived from a weighted score of over 30 indicators in seven different domains, including income, employment, health, education, skills and training, geographic access to services, crime, and housing [14, 15]. The SIMD is a relative measure of deprivation across 6,976 small areas termed data zones. SIMD quintile was recorded in the SMR01, and each quintile consisted of 20% of the data zones from the most deprived to the least deprived level [14].

### Statistical analyses

We estimated incidence of RSV-associated and influenza-associated RTI hospitalisations using two approaches, i.e., regression model-based approach and a direct measurement using ICD-10 diagnoses. The use of the two approaches allows us to understand the level of under-ascertainment of RSV across deprivation levels due to the lack of systematic RSV testing and poor sensitivity of RSV-specific ICD-10 codes in routine clinical care practice, and imperfect sensitivity of viral diagnostic tests.

For the model-based approach, we used a multiple linear regression model to estimate the average number of RTI hospitalisations associated with RSV (and influenza viruses) consistent with our recent analyses [11, 16, 17]. The model included a natural cubic spline function for weeks during the study period, the number of RSV-positive tests, and the number of influenza-positive tests. We considered a 0-3-week lag and/or lead for RSV and influenza in each model and tested for the optimal lag and/or lead combination for the two predictors simultaneously. Models were fitted separately by age group (0-2 months, 3-5 months, 6-11 months, 1-2 years, 3-4 years, 5-17 years, 18-64 years, 65-74 years, 75-84 years, and 85+ years). The goodness of fit was assessed based on an adjusted R-squared and Akaike Information Criterion (AIC). We estimated the annual number and rates of RSV-associated (and influenza-associated) RTI hospitalisations based on model coefficients for RSV (and influenza), the number of RSV-positive tests (and influenza-positive tests), and Scottish population statistics by SIMD and age [18]. The 95% confidence intervals (CIs) were estimated using a 52-week-block bootstrap with 1000 replicates.

For the direct measurement approach, we estimated RSV-associated (and influenza-associated) RTI hospitalisations based on ICD-10 diagnoses, by counting hospital episodes of ICD-10 coded RSV-associated (and influenza-associated) RTI [12]. Then we estimated annual rates of RSV-associated (and influenza-associated) RTI hospitalisation and 95% CIs from the Poisson distribution, based on Scottish population statistics by SIMD and age [18].

We then estimated rate ratios (RRs) of RSV-associated RTI hospitalisation and the 95% uncertainty range (UR) between the SIMD levels by age group. As previously done [19], the 95% UR of RR were derived using 1000 samples from log-normal distributions of RSV-associated RTI hospitalisation rates, with the 2.5^th^ percentile and the 97.5^th^ percentile as the lower and the upper bound.

### Sensitivity analyses

We conducted the following sensitivity analyses to assess the robustness of estimates of RSV-associated RTI hospitalisation: (1) using the negative binomial regression model; (2) adding an interaction term between influenza-positive tests and season (2010-11 season; other seasons) to the main models to account for potential differences in testing practices and influenza epidemiology in the 2010-11 season compared to other seasons; (3) adding time series data of rhinovirus-positive tests to the main models.

## Results

### Regression model-based estimates of RSV-associated RTI hospitalisation

From 2010 – 2016, the weekly RSV positive tests remained steady peaking around the same time each year whereas weekly influenza positive tests were highest in 2010 compared to other years (Figure S1). The weekly observed versus fitted RTI hospitalisations generally followed a similar pattern across SIMD levels. Time series of RTI hospitalisations and RSV positive tests are in the appendix (Figure S1).

Using the regression model-based approach, we estimated that the average annual number of RSV-associated RTI hospitalisations ranged from 884 for individuals of all ages in the least deprived group (5^th^ quintile by SIMD) to 1,676 for individuals in the most deprived group (1^st^ quintile by SIMD) (Table 1). Estimates of RSV-associated RTI hospitalisation rates gradually increased with levels of deprivation in individuals of all ages, with the highest rate of 1.51 (95% confidence interval (CI): 1.03 – 1.79) per 1 000 in the 1^st^ quintile by SIMD and lowest rate of 0.76 (0.43 – 0.93) per 1,000 in the 5^th^ quintile. A similar pattern of RSV-associated RTI hospitalisation rates with SIMD appeared to remain in most of the age groups, except in adults ≥85 years old. Across the SIMD and age groups, infants aged 0-2 months in the 1^st^ SIMD had the highest RSV-associated RTI hospitalisation rate of 75.77 (65.24 - 82.13) per 1,000 infants per year. Details on model structures by SIMD and age groups are in Supplementary Table 4.

**Table 1.** Model-based estimates of annual average number and rate of RSV-associated RTI hospital admission during 2010-2016, by age group and SIMD in Scotland.

|  | SIMD quintiles |  |  |  |  |
| --- | --- | --- | --- | --- | --- |
|  | 1st quintile | 2nd quintile | 3rd quintile | 4th quintile | 5th quintile |
| **Overall** |  |  |  |  |  |
| **Cases** | 1676 | 1368 | 1167 | 968 | 884 |
| **Rate (95% CI)** | 1.51 (1.03,1.79) | 1.22 (0.68,1.52) | 1.06 (0.72,1.26) | 0.86 (0.55,1.02) | 0.76 (0.43,0.9) |
| **0-2m** |  |  |  |  |  |
| **Cases** | 270 | 185 | 147 | 140 | 115 |
| **Rate (95% CI)** | 75.77 (65.24,82.13) | 60.07 (49.81,66.99) | 56.37 (47.07,62.47) | 52.37 (45.74,56.84) | 42.39 (33.54,46.85) |
| **3-5m** |  |  |  |  |  |
| **Cases** | 203 | 151 | 122 | 97 | 73 |
| **Rate (95% CI)** | 56.13 (47.83,62.25) | 49.8 (36.88,57.41) | 46.84 (40.28,51.01) | 36.56 (31.93,42.08) | 27.2 (22.9,30.16) |

\* SIMD: The Scottish Index of Multiple Deprivation is derived based on seven different domains, including income, employment, health, education, skills and training, geographic access to services, crime, and housing, with the 5th quintile indicating the least deprived groups in Scotland.

| SIMD quintiles |  |  |  |  |  |
| --- | --- | --- | --- | --- | --- |
|  | 1st quintile | 2nd quintile | 3rd quintile | 4th quintile | 5th quintile |
| **6-11m** |  |  |  |  |  |
| **Cases** | 211 | 156 | 116 | 123 | 93 |
| **Rate (95% CI)** | 29.49 (23.63,33.73) | 25.73 (18.97,31.03) | 21.29 (10.89,25.43) | 22.85 (17.01,26.77) | 17.24 (13.74,19.51) |
| **1-2y** |  |  |  |  |  |
| **Cases** | 327 | 222 | 184 | 177 | 164 |
| **Rate (95% CI)** | 11.56 (7.82,13.45) | 9.11 (6.1,10.81) | 8.34 (5.44,9.93) | 7.6 (5.02,10.05) | 7.07 (3.64,9.09) |
| **3-4y** |  |  |  |  |  |
| **Cases** | 78 | 94 | 61 | 45 | 43 |
| **Rate (95% CI)** | 2.63 (0.73,3.86) | 3.8(1.97,4.78) | 2.65 (0.98,3.23) | 1.82 (0.7,2.82) | 1.72 (0.72,2.38) |
| **5-17y** |  |  |  |  |  |
| **Cases** | 84 | 53 | 63 | 55 | 59 |
| **Rate (95% CI)** | 0.53 (0.27,0.75) | 0.31(0.09,0.46) | 0.44 (0.27,0.57) | 0.35 (0.15,0.48) | 0.33 (0.17,0.45) |
| **18-64y** |  |  |  |  |  |
|  | 1st quintile | 2nd quintile | 3rd quintile | 4th quintile | 5th quintile |
| **Cases** | 158 | 151 | 158 | 86 | 86 |
| **Rate (95% CI)** | 0.21 (-0.02,0.36) | 0.21(0.09,0.32) | 0.24(0.11,0.36) | 0.11(0.01,0.22) | 0.12 (0.06,0.18) |
| **65-74y** |  |  |  |  |  |
| **Cases** | 108 | 120 | 90 | 85 | 78 |
| **Rate (95% CI)** | 1.17 (0.45,1.78) | 1.08 (0.58,1.42) | 0.79 (0.24,1.09) | 0.71 (0.25,0.97) | 0.7 (0.3,0.9) |
| **75-84y** |  |  |  |  |  |
| **Cases** | 258 | 212 | 199 | 150 | 131 |
| **Rate (95% CI)** | 4.15 (2.37,5.35) | 2.94 (0.98,4.11) | 2.84(1.74,3.46) | 2.21 (0.96,2.72) | 1.92 (0.92,2.47) |
| **85y+** |  |  |  |  |  |
| **Cases** | 162 | 208 | 155 | 167 | 200 |
| **Rate (95% CI)** | 7.07 (1.6,9.86) | 8.06(4.71,10.68) | 5.52 (1.43,7.84) | 6.78 (2.44,8.9) | 8.22 (4.92,10.42) |

### Estimates of ICD-10 coded RSV-associated RTI hospitalisation

We found lower average annual number and rate of ICD-10 coded RSV-associated RTI hospitalisations compared to the model-based estimates, across SIMD and age groups (Tables 1 and 2). Similar to the model-based estimates, RSV-RTI hospitalisation estimates based on ICD-10 diagnoses generally increased with levels of deprivation. As shown in Table 2, we estimated that the average annual number of RSV-coded RTI hospitalisations ranged from 242 for individuals of all ages in the 5^th^ quintile of SIMD to 498 for individuals in the 1^st^ quintile of SIMD. Individuals in the 1^st^ quintile and 5^th^ quintile of SIMD had the highest and lowest RSV-coded RTI hospitalisation rate, at 0.47 (95% CI: 0.46 – 0.49) and 0.22 (95% CI: 0.21 – 0.23) per 1,000. By age groups, a similar decreasing pattern of RSV-associated RTI hospitalisation rates with SIMD was mainly seen in children under 2 years old. Estimates of RSV-associated RTI hospitalisations in children above 5 years old were mostly too low to allow for comparison, roughly between 0.01 and 0.07 (95% CI: 0.04 – 0.13) per 1,000. Using this approach, infants aged 0-2 months in the 1^st^ quintile of SIMD, among all the SIMD and age groups, had the highest RSV-associated RTI hospitalisation rate of 45.79 (95% CI: 42.96 – 48.77) per 1,000 per year.

**Table 2.** Estimates of annual average number and rate of ICD-10 coded RSV-associated RTI hospital admission during 2010-2016, by age group and SIMD in Scotland.

|  | SIMD quintiles |  |  |  |  |
| --- | --- | --- | --- | --- | --- |
|  | 1st quintile | 2nd quintile | 3rd quintile | 4th quintile | 5th quintile |
| **Overall** |  |  |  |  |  |
| **Cases** | 498 | 378 | 304 | 290 | 242 |
| **Rate (95% CI)** | 0.47 (0.46,0.49) | 0.36 (0.34,0.37) | 0.29 (0.27,0.3) | 0.27 (0.26,0.28) | 0.22 (0.21,0.23) |
| **0-2m** |  |  |  |  |  |
| **Cases** | 162 | 120 | 98 | 91 | 72 |
| **Rate (95% CI)** | 45.79 (42.96,48.77) | 40.03 (37.16,43.07) | 37.29 (34.33,40.43) | 34.6 (31.76,37.62) | 27.65 (25.1,30.38) |
| **3-5m** |  |  |  |  |  |
| **Cases** | 110 | 87 | 66 | 52 | 42 |
| **Rate (95% CI)** | 30.98 (28.66,33.45) | 29.06 (26.62,31.67) | 25.01 (22.59,27.6) | 19.76 (17.63,22.07) | 15.94 (14.02,18.04) |

<sup>2</sup> SIMD: The Scottish Index of Multiple Deprivation is derived based on seven different domains, including income, employment, health, education, skills and training, geographic access to services, crime, and housing, with the 5th quintile indicating the least deprived groups in Scotland.

| SIMD quintiles |  |  |  |  |  |
| --- | --- | --- | --- | --- | --- |
|  | 1st quintile | 2nd quintile | 3rd quintile | 4th quintile | 5th quintile |
| **6-11m** |  |  |  |  |  |
| **Cases** | 103 | 77 | 63 | 62 | 52 |
| **Rate (95% CI)** | 14.62 (13.49,15.82) | 12.81 (11.66,14.03) | 12.03(10.84,13.3) | 11.81 (10.64,13.06) | 10.05 (8.97,11.22) |
| **1-2y** |  |  |  |  |  |
| **Cases** | 102 | 75 | 59 | 67 | 60 |
| **Rate (95% CI)** | 3.66 (3.38,3.96) | 3.14 (2.86,3.45) | 2.79 (2.51,3.09) | 3.06 (2.77,3.38) | 2.73 (2.46,3.03) |
| **3-4y** |  |  |  |  |  |
| **Cases** | 7 | 6 | 5 | 6 | 7 |
| **Rate (95% CI)** | 0.24 (0.17,0.33) | 0.25 (0.17,0.35) | 0.24 (0.16,0.34) | 0.28 (0.19,0.38) | 0.31 (0.23,0.42) |
| **5-17y** |  |  |  |  |  |
| **Cases** | 4 | 4 | 5 | 3 | 2 |
| **Rate (95% CI)** | 0.02 (0.01,0.04) | 0.03 (0.02,0.04) | 0.04 (0.03,0.05) | 0.02 (0.01,0.03) | 0.01 (0.01,0.02) |
| **18-64y** |  |  |  |  |  |
|  | 1st quintile | 2nd quintile | 3rd quintile | 4th quintile | 5th quintile |
| **Cases** | 6 | 5 | 4 | 4 | 3 |
| **Rate (95% CI)** | 0.01 (0.01,0.01) | 0.01 (0,0.01) | 0.01 (0,0.01) | 0.01 (0,0.01) | 0 (0,0.01) |
| **65-74y** |  |  |  |  |  |
| **Cases** | 2 | 2 | 2 | 2 | 2 |
| **Rate (95% CI)** | 0.02 (0.01,0.04) | 0.02 (0.01,0.04) | 0.02 (0.01,0.03) | 0.02 (0.01,0.04) | 0.02 (0.01,0.03) |
| **75-84y** |  |  |  |  |  |
| **Cases** | 3 | 3 | 2 | 2 | 2 |
| **Rate (95% CI)** | 0.04 (0.02,0.07) | 0.05 (0.03,0.07) | 0.03 (0.02,0.05) | 0.03 (0.02,0.06) | 0.03 (0.01,0.05) |
| **85y+** |  |  |  |  |  |
| **Cases** | 0 | 1 | 2 | 1 | 2 |
| **Rate (95% CI)** | 0.02 (0,0.06) | 0.03 (0.01,0.07) | 0.07 (0.03,0.13) | 0.05 (0.02,0.11) | 0.07 (0.04,0.13) |

### Estimates of influenza-associated RTI hospitalisation by SIMD and age group

The highest estimated average annual number and rate of influenza-associated RTI hospitalisation in the younger age groups were observed in the 4^th^ quintile (Supplementary Table 2). Compared to other SIMD levels, the estimates were higher among age groups 0-2 months (140 cases; 52.37, 95% CI: 45.74 – 56.84); 3-5 months (97 cases; 36.56, 95% CI: 31.93 – 42.08); 6-11 months (123 cases; 22.85, 95% CI: 17.01 – 26.77); 1-2 years (177 cases; 7.6, 95% CI: 5.02 – 10.05); and 3-4 years (45 cases; 0.35, 95% CI: 0.15 – 0.48) in the 4^th^ quintile.

In adult age groups, the highest number of influenza-associated RTI hospitalisation was estimated in the 1^st^ quintile among 18-64 years – 408 cases at a rate of 0.57 (95% CI: 0.43 – 0.66) per 1,000, however, the rate of hospitalisation was higher (4.4, 95% CI: 1.96 – 12.89) per 1,000 among adults aged ≥85 years in the 1^st^ quintile. Similarly, the admission rates were higher among adults aged ≥85 years compared to other adult age groups across the SIMD levels (Supplementary Table 2).

### The ratio of RSV-RTI hospitalisation rates between SIMD by age group

The two approaches, i.e., ICD-10-based and model-based approach, generally yielded comparable RRs of RSV-RTI hospitalisations between SIMD levels, except in individuals aged 5-17 years old and ≥85 years old (Figure 1). Compared to the 1^st^ quintile of SIMD, the RR estimates showed an increasing pattern with higher deprivation levels in individuals of all ages (Figure 1, Supplementary Table 3). In detail, the RR was 2.13 (95% CI: 2.0 – 2.29), 1.62 (95% CI: 1.52-1.74), 1.29 (95% CI: 1.21-1.39) and 1.22 (95% CI: 1.15-1.32) in the 1^st^ to 4^th^ quintile of SIMD based on the ICD-10 approach, and 1.96 (95% CI: 1.23 – 3.25), 1.60 (95% CI: 1.0-2.66), 1.35 (95% CI: 0.85-2.25) and 1.12 (95% CI: 0.7-1.85) using the model-based approach. By age groups, the RR for the 1^st^ quintile of SIMD ranged from 0.24 (95% CI: 0.10 – 0.60) in adults aged ≥85 years to 2.33 (95% CI: 1.22 – 4.69) in adults aged 18-64 years based on the ICD-10 approach (Supplementary Table 3). The RR for the 1^st^ quintile of SIMD ranged from 0.96 (95% CI: 0.60 – 1.62) in adults aged ≥85 years to 2.08 (95% CI: 1.11 – 4.13) in adults aged 75-84 years (Supplementary Table 3) using the model-based approach. Individuals in the 1^st^ quintile of SIMD had the highest RR in children aged 2 years old and below. By contrast, the RR estimates by SIMD overlapped 1 on most of the occasions and were close to 1 (either above or below 1) on several occasions in people of 3-84 years old, suggesting no apparent patterns associated with SIMD in these age groups. Lastly, the RR in adults aged ≥85 years old was 0.24 (95% CI: 0.1-0.6) and 0.4 (95% CI: 0.17 - 1.0) in the 1^st^ and 2^nd^ quintile of SIMD by ICD-10 approach, while it overlapped 1 using the model-based approach (Supplementary Table 3).

**Figure 1.**
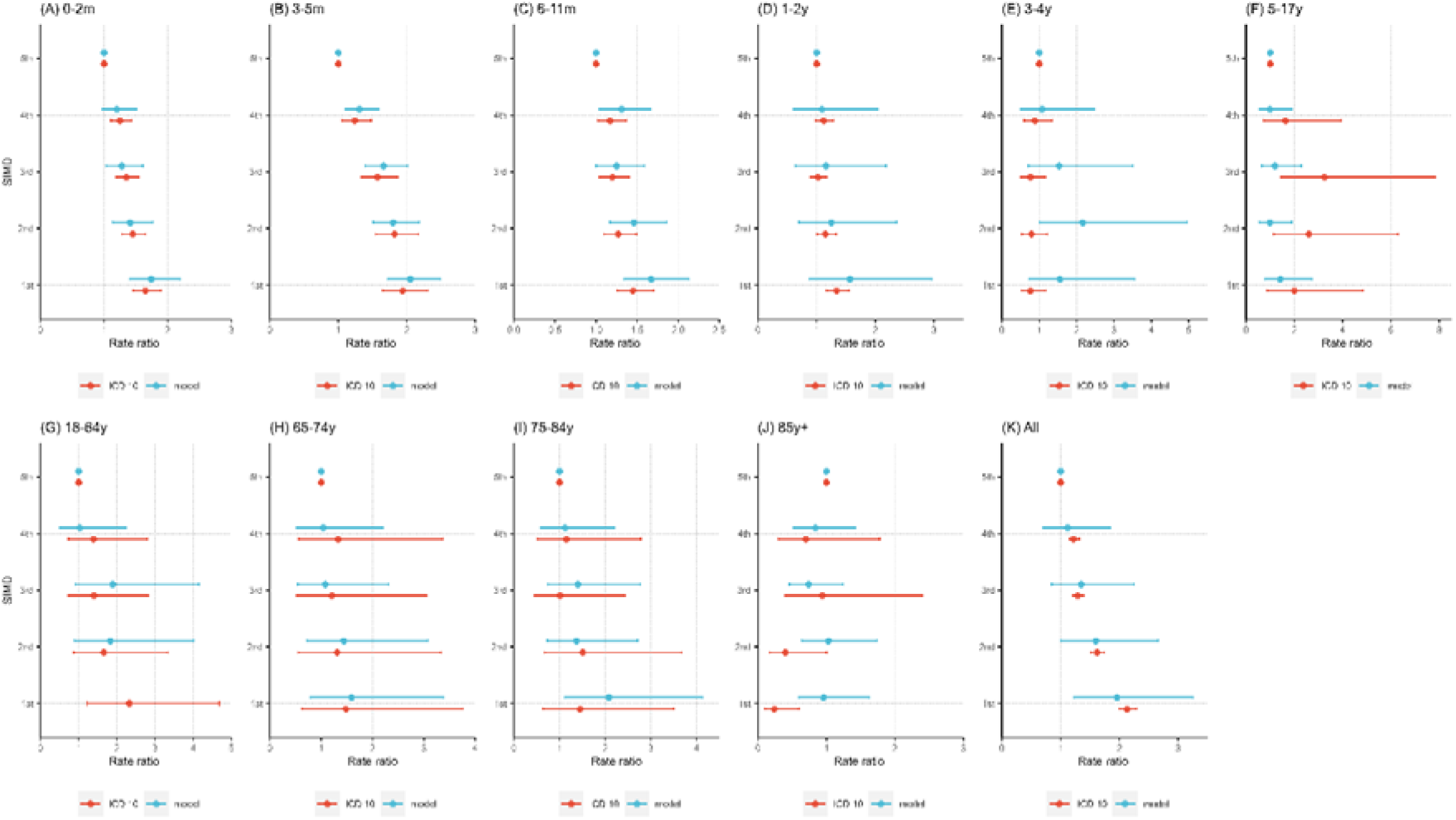
Ratio of RSV-RTI hospitalisation rates between SIMD quintiles, by age group. Panels show rate ratios of population of other SIMD levels compared with the least deprived (i.e., 5^th^ quintile of SIMD) by age group, i.e., 0-2 months (A), 3-5 months (B), 6-11 months (C), 1-2 years (D), 3-4 years (E), 5-17 years (F), 18-64 years (G), 65-74 years (H), 75-84 years (I), ≥85 years (J), and all the population (K).

### Sensitivity analyses

Estimates from all the sensitivity analyses are presented in Supplementary Table 5. In general, rates of RSV-RTI hospitalisations were comparable across SIMD levels when considering the main models and the sensitivity analyses. The use of negative binomial regression model and addition of rhinovirus yielded higher AIC values compared with models used in the main analyses.

## Discussion

Using two approaches on data from the Scottish national healthcare data and virological surveillance, we found that the rate of RSV-associated hospitalisation is generally higher among individuals in the most deprived groups (1^st^ quintile) compared to the least deprived groups (5^th^ quintile) in Scotland. In the general population, we found the highest average annual number of RSV-associated RTI hospitalisation and rate of admission in individuals in the most deprived group. The rate of RSV-associated RTI hospitalisation in the most deprived group was about twice as high as the rate of admission in the least deprived group. The differences in hospitalisation rates were most pronounced in infants and children aged 1-2 years old. Our analysis found that the rates of RSV-associated hospitalisation in children less than 1 year were up to about twice as high in the most deprived groups compared to the least deprived groups. This observation could be related to previously reported risk factors for respiratory infection transmission – family size, crowding, smoking, exposure to industrial pollutants and inadequate hygiene that are more prevalent among socioeconomically deprived groups [9, 20, 21].

The relationship with deprivation level was strongest in infants and to a lesser extent young children 1-2 years of age. In contrast, the patterns of RSV-associated RTI hospitalisation rates with SIMD in older children and adults showed a similar trend but this was less clear. There was no relationship observed in the oldest age group of those ≥85 years of age. Possible explanations for this may be age-group specific and may include for instance relatively small numbers and rates of RSV-associated RTI hospitalisations and confounding effects of other factors distributed between SIMD for 3-4 years old. In adults aged ≥85 years, differences by diagnosis and coding practice between SIMD, and confounding effect of other factors distributed between SIMD may explain this as we estimated RR of 2.0 (95% CI: 0.88 - 4.84) in the 1^st^ quintile of SIMD compared with the 5^th^ quintile of SIMD using the ICD-10 approach.

However, the highest average annual number of RSV-associated RTI hospitalisations (258 cases) in the adult population was seen among those aged 75-84 years in the most deprived group. In the adult age groups in general, though the admission rates varied minimally across the SIMD levels, no large variations were observed suggesting that age, in addition to deprivation level, is a significant determinant of RSV-associated RTI hospitalisations. In the elderly, other co-existing risk factors such as chronic medical conditions may play a more critical role in the risk profiles for RSV hospitalisations.

We observed that unlike the RSV hospitalisation pattern, the rates of influenza-associated RTI hospitalisation were higher in less deprived group (4^th^ quintile) compared to other SIMD levels across age groups for children less than 5 years (Supplementary Table 2). Among adults, the rates of influenza-associated hospitalisation were higher in those aged ≥85 years in the most deprived group. When considering the model-based and ICD-10 RR estimates of RSV-associated RTI hospitalisation, we observed a consistently higher risk of admission for children aged 0-2 years in the most deprived groups compared to the least deprived group while the RR remained similar across the SIMD levels for children aged 3-4 years. Our study period included the 2009 H1N1 pandemic where influenza-associated RTI hospitalisations were potentially disproportionately higher in the less deprived individuals and groups. Previous research reports that low social class was one of the factors associated with the risk of hospitalisation in children with bronchiolitis [22], and children from lower socioeconomic groups were at increased risk of admission to paediatric intensive care for bronchiolitis [23]. Studies suggest that the transmission of RSV may differ due to socially patterned risk factors such as residential overcrowding and family characteristics, which may lead to different patterns of hospitalisations [20, 21].

The SIMD measure is widely used to describe and assess Scottish small area concentrations of deprivation; however, reports suggest that individuals in certain areas could be missed [24, 25]. For instance, individuals experiencing deprivation may be more dispersed in rural areas, which may lead to greater heterogeneity in this population. The SIMD tends to privilege urban areas of deprivation compared to deprived individuals in more rural areas [24]. It is more sensitive at detecting income and employment deprived individuals in urban areas compared to remote and rural areas and island local authorities [25]. The percentage of income and employment deprived individuals missed by the SIMD is greater in remote and rural areas, however, the absolute number of people missed is higher in urban areas due to higher deprivation levels [25].

In this study, we did not have access to the sub-domains to explore which domains contribute to the effects we observed. The lower estimates from the ICD-10 direct measurement approach compared to the modelling approach may be explained by the limitations of using ICD-10 codes without laboratory confirmation in respiratory disease classification/diagnosis, especially in adults.

Our study highlights the burden of RSV-associated hospitalisation in the overall population and by age group across SIMD levels and demonstrates that children in the most deprived groups may be suffering a higher burden of RSV hospitalisations. It further shows that RSV hospitalisation in older adults across deprivation levels may be similar. In addition to the age-specific vulnerability of RSV hospitalisation for children less than 5 years, being in a deprived group may present higher risks of RSV-associated RTI hospitalisation. Our study highlights the need to target children in low socioeconomic groups or in the most deprived groups for any future prevention strategies and interventions especially as RSV vaccines become available.

## Conclusion

Our analysis focused on estimating RSV and influenza-associated hospitalisation in children and adults based on socioeconomic status using SIMD levels in Scotland. Our results show that RSV hospitalisation rates are about twice as much in groups that are most deprived compared to the least deprived group in Scotland. We observed that the deprivation-related disparity in RSV hospitalisation rates were more pronounced in children of 2 years old and below than in other age groups. These results underscore the need to create more awareness of RSV-associated hospitalisation among individuals in deprived groups and areas at various levels in the hospital/clinical setup, and it may also be useful to consider this as part of the triage and/or treatment strategy. The results also highlight the importance of prioritising individuals in deprived areas for future interventions and RSV prevention strategies.

## Data Availability

All data produced in the present work are contained in the manuscript. Request for these data may be made to the electronic data research and innovation services (eDRIS) team at Public Health Scotland

## Footnote Page

The PROMISE investigators are as follows:

Harish Nair (University of Edinburgh), Hanna Nohynek (THL), Terho Heikkinen (University of Turku and Turku University Hospital), Anne Teirlinck (RIVM), Louis Bont (University Medical Center Utrecht), Philippe Beutels (University of Antwerp), Peter Openshaw (Imperial College, London), Andrew Pollard (University of Oxford), Alexandro Orrico Sánchez (FISABIO), Veena Kumar (Novavax), Tin Tin Htar (Pfizer), Charlotte Vernhes (Sanofi Pasteur), Gael Dos Santos (GlaxoSmithKline), Jeroen Aerssens (Janssen), Rolf Kramer (Sanofi Pasteur), Nuria Manchin (TEAMIT).

## Acknowledgements

We acknowledge the support of the electronic data research and innovation services (eDRIS) team at Public Health Scotland for their involvement in obtaining approvals, provisioning and linking, and the use of the secure analytical platform with the National Safe Haven.

