## Supplementary File for "Respiratory syncytial virus-associated hospital admissions by deprivation levels among children and adults in Scotland"

**Supplementary file 1**

Model structure

We used the multiple linear regression model to estimate the number of RTI hospital admissions associated with RSV and influenza viruses in children and adults based by SIMD level and age group as previously done [1-3]. Overall, the model included a natural cubic spline function for weeks during the study period, the number of RSV-positive tests, and the number of influenza-positive tests.

$$E(Y_{t,a})={\beta_{1,a}S}_{t}+\beta_{2,a}RSV_{t-ta1}+\beta_{3,a}Flu_{t-ta2}$$

t: week index (from 1 to the length of the study period)

a: Age group a

Y: weekly number of RTI hospital episodes (3-week moving average)

S: a natural cubic spline function of weeks.

RSV: weekly RSV positive tests

Flu: influenza-positive tests.

ta1: lags and/or leads in RSV

ta2: lags and/or leads in influenza.

**Supplementary file 2**

Supplementary Table 1: The list of ICD codes used to identify relevant diagnosis group (RED is other-pathogen code, GREEN is RSV code, BLUE is influenza code) [4]

| **Diagnosis** | | **ICD-10** |
| --- | --- | --- |
| Acute upper respiratory tract infection (URTI) | | J00 **J02.0 J02.8** J02.9 **J03.0 J03.8** J03.9 J04.0 J04.1 J04.2 J05.0 J05.1 J06.0 J06.8 J06.9 |
| Lower respiratory tract infection (LRTI) | Pneumonia and influenza | **J09 J10.0 J10.1 J10.8 J11.0 J11.1 J11.8** **J12.0** **J12.1** **J12.2** J12.3 J12.8 J12.9 **J13 J14 J15.0 J15.1 J15.2 J15.3 J15.4 J15.5 J15.6 J15.7 J15.8** J15.9 **J16.0 J16.8 J17.0 J17.1 J17.2 J17.3 J17.8** J18.0 J18.1 J18.2 J18.8 J18.9 |
|  | Bronchiolitis and bronchitis | **J20.0 J20.1 J20.2 J20.3 J20.4** **J20.5** **J20.6 J20.7 J20.8** J20.9 **J21.0** **J21.1 J21.8** J21.9 J40 |
|  | Unspecified LRTI | J22 |
| Pathogen-specific codes** (only included when one of above J codes is also used) | | B95: Streptococcus, Staphylococcus**: B95.0 B95.1 B95.2 B95.3 B95.4 B95.5 B95.6 B95.7 B95.8**  B96: other bacterial agents: **B96.0 B96.1 B96.2 B96.3 B96.4 B96.5 B96.6 B96.7 B96.8**  B97: viral agents: **B97.0 B97.1 B97.2 B97.3 B97.4 B97.5 B97.6 B97.7 B97.8**  B98: other specified infectious agents: **B98.0 B98.1** |

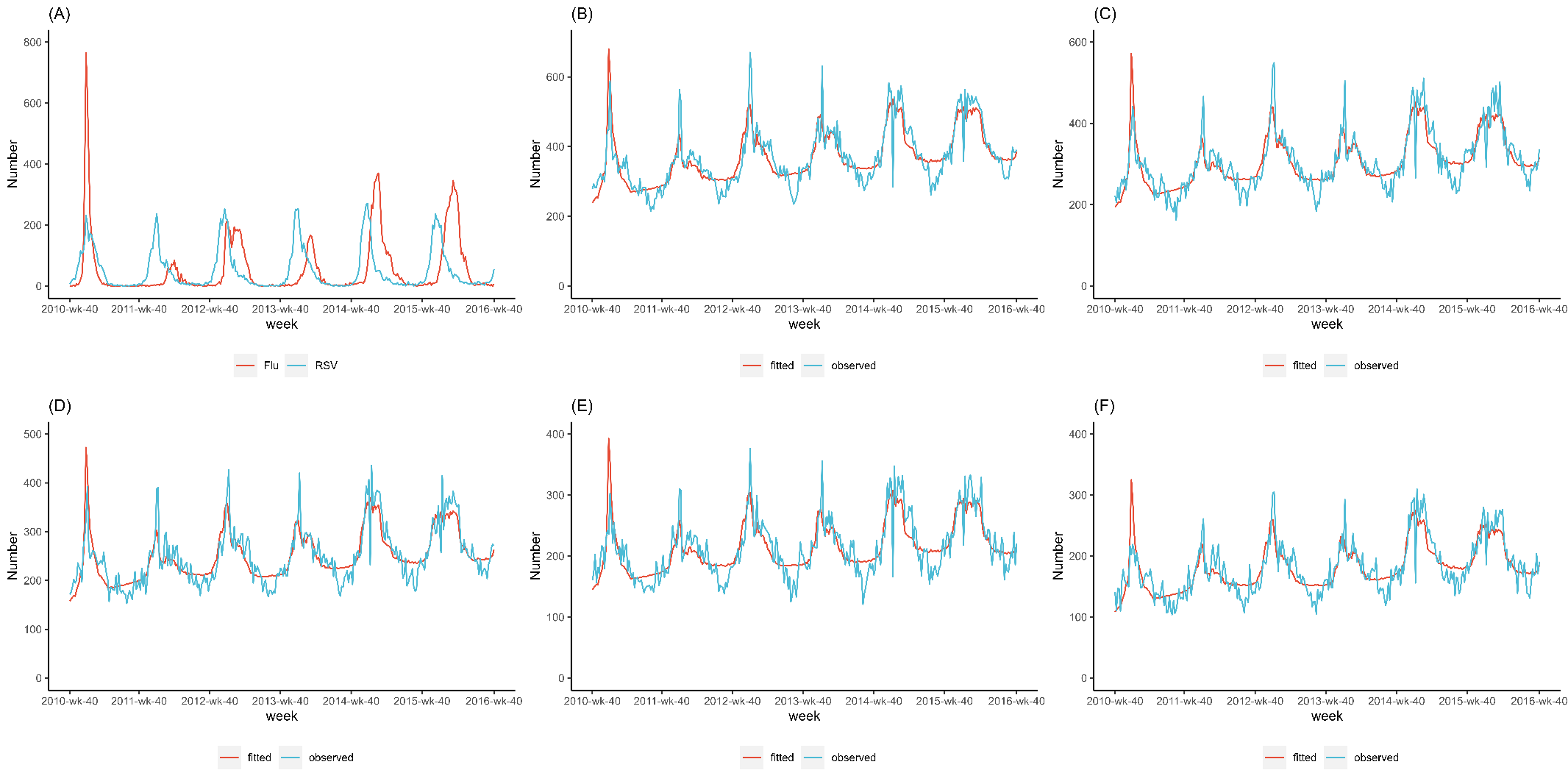

**Figure S1.** Weekly time series for respiratory syncytial virus (RSV) and influenza (A), and weekly time series of observed and fitted respiratory tract infection (RTI) hospital admissions by SIMD (Panel B-F). Panel B to F shows the observed RTI cases and fitted RTI cases in population of the 1^st^, 2^nd^, 3^rd^, 4^th^, and the 5^th^ quintile of SIMD.

**Supplementary Table 2.** Model-based estimates of influenza-associated RTI hospital admission during 2010-2016, by age group and SIMD in Scotland.

|  |  | Age-groups | | | | | | | | | |
| --- | --- | --- | --- | --- | --- | --- | --- | --- | --- | --- | --- |
| SIMD^a^ | **Estimate** | **0-2m** | **3-5m** | **6-11m** | **1-2y** | **3-4y** | **5-17y** | **18-64y** | **65-74y** | **75-84y** | **≥85y** |
| 1st quintile | Hospital admissions | 8 | 20 | 33 | 71 | 43 | 76 | 408 | 148 | 157 | 98 |
|  | Hospital admission rate per 1000 population (95% CI) | 1.39  (-1.9,10.97) | 3.64  (-2.08,13.79) | 3.47  (-1.14,10.8) | 2.16  (0.38,4.87) | 1.32  (0.33,3.27) | 0.4  (0.27,0.72) | 0.57  (0.43,0.66) | 1.46  (0.89,2.3) | 2.45  (1.22,4.39) | 4.4  (1.96,12.89) |
| 2nd quintile | Hospital admissions | 11 | 13 | 21 | 77 | 27 | 57 | 330 | 113 | 138 | 103 |
|  | Hospital admission rate per 1000 population (95% CI) | 3.09  (-2.27,9.09) | 3.04  (-1.57,10.99) | 2.48  (-1.12,8.01) | 2.67  (0.65,5.19) | 0.94  (-0.01,2.41) | 0.36  (0.28,0.54) | 0.47  (0.38,0.64) | 0.99  (0.44,1.67) | 1.89  (0.91,3.85) | 3.91  (1.66,10.56) |
| 3rd quintile | Hospital admissions | 20 | 16 | 23 | 46 | 32 | 49 | 256 | 100 | 97 | 115 |
|  | Hospital admission rate per 1000 population (95% CI) | 7.14  (2.8,14.18) | 4.68  (1.07,9.22) | 3.41  (-1.63,10.3) | 1.79  (-0.02,4.14) | 1.3  (0.63,2.01) | 0.34  (0.12,0.51) | 0.36  (0.26,0.45) | 0.78  (0.41,1.28) | 1.2  (0.31,3.01) | 4.23  (1.65,9.29) |
| 4th quintile | Hospital admissions | 140 | 97 | 123 | 177 | 45 | 55 | 86 | 85 | 150 | 167 |
|  | Hospital admission rate per 1000 population (95% CI) | 52.37  (45.74,56.84) | 36.56  (31.93,42.08) | 22.85  (17.01,26.77) | 7.6  (5.02,10.05) | 1.82  (0.7,2.82) | 0.35  (0.15,0.48) | 0.11  (0.01,0.22) | 0.71  (0.25,0.97) | 2.21  (0.96,2.72) | 6.78  (2.44,8.9) |
| 5th quintile | Hospital admissions | -1 | 9 | 2 | 40 | 25 | 39 | 154 | 57 | 98 | 72 |
|  | Hospital admission rate per 1000 population (95% CI) | -1.23  (-7.14,2.35) | 3.52  (0.12,8.55) | 0.12  (-2,3.39) | 1.48  (0.36,4.04) | 0.83  (-0.02,2.13) | 0.23  (0.12,0.4) | 0.22  (0.16,0.27) | 0.48  (0.23,0.83) | 1.38  (0.55,3.36) | 2.58  (0.25,5.94) |

a: The Scottish Index of Multiple Deprivation is derived based on seven different domains, including income, employment, health, education, skills and training, geographic access to services, crime, and housing, with the 5^th^ quintile indicating the least deprived groups in Scotland.

**Supplementary Table 3.** Ratio of RSV-RTI hospital admission rates between SIMD groups, by age group.

|  | **Scottish Index of Multiple Deprivation** | | | | | |
| --- | --- | --- | --- | --- | --- | --- |
|  | | **1st quintile** | **2nd quintile** | **3rd quintile** | **4th quintile** | **5th quintile** |
| **Overall** | |  |  |  |  |  |
| **RR (95% CI) based on ICD-10** | | 2.13 (2.0-2.29) | 1.62 (1.52-1.74) | 1.29 (1.21-1.39) | 1.22 (1.15-1.32) | Reference |
| **RR (95% CI) based on Model** | | 1.96 (1.23-3.25) | 1.60 (1.0-2.66) | 1.35 (0.85-2.25) | 1.12 (0.7-1.85) | Reference |
| **0-2m** | |  |  |  |  |  |
| **RR (95% CI) based on ICD-10** | | 1.65 (1.46-1.89) | 1.45 (1.28-1.65) | 1.35 (1.19-1.54) | 1.25 (1.11-1.43) | Reference |
| **RR (95% CI) based on Model** | | 1.74 (1.4-2.19) | 1.41 (1.13-1.77) | 1.28 (1.03-1.61) | 1.20 (0.97-1.51) | Reference |
| **3-5m** | |  |  |  |  |  |
| **RR (95% CI) based on ICD-10** | | 1.94 (1.65-2.31) | 1.82 (1.55-2.17) | 1.57 (1.33-1.87) | 1.24 (1.05-1.48) | Reference |
| **RR (95% CI) based on Model** | | 2.05 (1.72-2.49) | 1.80 (1.51-2.18) | 1.66 (1.39-2.01) | 1.31 (1.1-1.59) | Reference |
| **6-11m** | |  |  |  |  |  |
| **RR (95% CI) based on ICD-10** | | 1.45 (1.26-1.70) | 1.27 (1.1-1.49) | 1.20 (1.04-1.40) | 1.17 (1.02-1.37) | Reference |
| **RR (95% CI) based on Model** | | 1.67 (1.34-2.13) | 1.46 (1.17-1.86) | 1.25 (1.0-1.59) | 1.31 (1.04-1.66) | Reference |
| **1-2y** | |  |  |  |  |  |
| **RR (95% CI) based on ICD-10** | | 1.34 (1.17-1.55) | 1.15 (1.01-1.33) | 1.02 (0.89-1.18) | 1.12 (0.98-1.29) | Reference |
| **RR (95% CI) based on Model** | | 1.57 (0.87-2.96) | 1.25 (0.7-2.36) | 1.16 (0.64-2.18) | 1.09 (0.61-2.05) | Reference |
| **3-4y** | |  |  |  |  |  |
| **RR (95% CI) based on ICD-10** | | 0.76 (0.52-1.17) | 0.79 (0.53-1.21) | 0.76 (0.51-1.17) | 0.88 (0.59-1.34) | Reference |
| **RR (95% CI) based on Model** | | 1.55 (0.72-3.55) | 2.16 (1.0-4.96) | 1.53 (0.71-3.5) | 1.08 (0.5-2.47) | Reference |
| **5-17y** | |  |  |  |  |  |
| **RR (95% CI) based on ICD-10** | | 2.0 (0.88-4.84) | 2.6 (1.15-6.29) | 3.24 (1.43-7.85) | 1.63 (0.72-3.94) | Reference |
| **RR (95% CI) based on Model** | | 1.42 (0.78-2.73) | 0.99 (0.54-1.89) | 1.20 (0.65-2.29) | 0.99 (0.54-1.89) | Reference |
| **18-64y** | |  |  |  |  |  |
| **RR (95% CI) based on ICD-10** | | 2.33 (1.22-4.69) | 1.66 (0.87-3.34) | 1.40 (0.73-2.82) | 1.39 (0.73-2.8) | Reference |
| **RR (95% CI) based on Model** | | No filled | 1.83 (0.89-4.01) | 1.89 (0.91-4.15) | 1.03 (0.5-2.25) | Reference |
| **65-74y** | |  |  |  |  |  |
| **RR (95% CI) based on ICD-10** | | 1.48 (0.63-3.76) | 1.31 (0.55-3.33) | 1.21 (0.51-3.06) | 1.33 (0.56-3.37) | Reference |
| **RR (95% CI) based on Model** | | 1.59 (0.79-3.38) | 1.44 (0.72-3.08) | 1.08 (0.54-2.31) | 1.04 (0.51-2.21) | Reference |
| **75-84y** | |  |  |  |  |  |
| **RR (95% CI) based on ICD-10** | | 1.45 (0.64-3.5) | 1.51 (0.67-3.67) | 1.01 (0.44-2.44) | 1.15 (0.51-2.78) | Reference |
| **RR (95% CI) based on Model** | | 2.08 (1.11-4.13) | 1.37 (0.73-2.71) | 1.40 (0.74-2.77) | 1.12 (0.59-2.21) | Reference |
| **≥85y** | |  |  |  |  |  |
| **RR (95% CI) based on ICD-10** | | 0.24 (0.10-0.60) | 0.4 (0.17-1.0) | 0.94 (0.4-2.4) | 0.7 (0.3-1.78) | Reference |
| **RR (95% CI) based on Model** | | 0.96 (0.60-1.62) | 1.03 (0.64-1.74) | 0.74 (0.46-1.24) | 0.84 (0.52-1.42) | Reference |

**Supplementary Table 4.** Model structure by age group and SIMD.

|  |  | Age-groups | | | | | | | | | |
| --- | --- | --- | --- | --- | --- | --- | --- | --- | --- | --- | --- |
| SIMD^a^ | **Term** | **0-2m** | **3-5m** | **6-11m** | **1-2y** | **3-4y** | **5-17y** | **18-64y** | **65-74y** | **75-84y** | **≥85y** |
| 1st quintile | Model AIC^b^ | 1483.25 | 1462.19 | 1864.69 | 1910.80 | 1615.86 | 1711.09 | 2326.46 | 2015.35 | 2104.65 | 1994.11 |
|  | Lag combination^c^ | (0,3) | (0,2) | (0,3) | (-1,2) | (-2,3) | (-3,3) | (3,0) | (3,1) | (3,0) | (3,0) |
|  | Coefficient of RSV (95%CI) | 0.1  (0.1-0.1) | 0.07  (0.07-0.08) | 0.07  (0.08-0.09) | 0.11  (0.12-0.13) | 0.02  (0.03-0.03) | 0.02  (0.03-0.04) | 0.04  (0.06-0.08) | 0.03  (0.04-0.05) | 0.08  (0.1-0.11) | 0.05  (0.06-0.07) |
|  | Coefficient of influenza | 0 | 0.01 | 0.01 | 0.02 | 0.02 | 0.03 | 0.14 | 0.05 | 0.05 | 0.03 |
| 2nd quintile | Model AIC^b^ | 1339.63 | 1301.78 | 1721.97 | 1837.34 | 1512.37 | 1611.67 | 2135.25 | 1923.75 | 2116.62 | 1969.28 |
|  | Lag combination^c^ | (1,3) | (0,3) | (0,3) | (-2,1) | (-1,3) | (-2,3) | (2,0) | (3,1) | (3,1) | (3,0) |
|  | Coefficient of RSV (95%CI) | 0.07  (0.07-0.07) | 0.05  (0.06-0.06) | 0.05  (0.06-0.06) | 0.07  (0.08-0.09) | 0.03  (0.03-0.04) | 0.01  (0.02-0.03) | 0.04  (0.06-0.07) | 0.04  (0.04-0.05) | 0.07  (0.08-0.09) | 0.07  (0.08-0.09) |
|  | Coefficient of influenza | 0 | 0 | 0.01 | 0.03 | 0.01 | 0.02 | 0.11 | 0.04 | 0.05 | 0.04 |
| 3rd quintile | Model AIC^b^ | 1161.82 | 1082.85 | 1678.95 | 1687.64 | 1383.85 | 1530.48 | 2085.82 | 1830.88 | 1999.78 | 1979.61 |
|  | Lag combination^c^ | (0,-1) | (0,3) | (0,3) | (-1,3) | (-1,3) | (-2,2) | (1,0) | (3,0) | (3,1) | (3,0) |
|  | Coefficient of RSV (95%CI) | 0.05  (0.05-0.06) | 0.04  (0.04-0.05) | 0.04  (0.04-0.05) | 0.06  (0.07-0.07) | 0.02  (0.02-0.03) | 0.02  (0.02-0.03) | 0.05  (0.06-0.07) | 0.03  (0.03-0.04) | 0.06  (0.07-0.08) | 0.05  (0.06-0.07) |
|  | Coefficient of influenza | 0.01 | 0.01 | 0.01 | 0.02 | 0.01 | 0.02 | 0.09 | 0.03 | 0.03 | 0.04 |
| 4th quintile | Model AIC^b^ | 1184.57 | 1034.86 | 1642.67 | 1717.34 | 1404.94 | 1509.99 | 1977.94 | 1723.18 | 1919.45 | 1909.26 |
|  | Lag combination^c^ | (1,-3) | (0,2) | (-1,3) | (-2,3) | (-2,3) | (-2,3) | (3,0) | (3,1) | (3,1) | (3,-3) |
|  | Coefficient of RSV (95%CI) | 0.05  (0.05-0.05) | 0.03  (0.04-0.04) | 0.04  (0.05-0.05) | 0.06  (0.07-0.07) | 0.01  (0.02-0.02) | 0.02  (0.02-0.03) | 0.02  (0.03-0.04) | 0.0  2(0.03-0.04) | 0.05  (0.06-0.07) | 0.05  (0.06-0.07) |
|  | Coefficient of influenza | 0 | 0 | 0.01 | 0.02 | 0.01 | 0.01 | 0.07 | 0.03 | 0.03 | 0.02 |
| 5th quintile | Model AIC^b^ | 1108.77 | 989.84 | 1525.90 | 1685.79 | 1254.50 | 1424.98 | 1767.25 | 1604.99 | 1893.03 | 1871.81 |
|  | Lag combination^c^ | (0,-3) | (0,3) | (-1,3) | (-2,3) | (-2,3) | (-2,3) | (3,0) | (3,1) | (3,0) | (2,-1) |
|  | Coefficient of RSV (95%CI) | 0.04  (0.04-0.04) | 0.02  (0.03-0.03) | 0.03  (0.03-0.04) | 0.05  (0.06-0.07) | 0.01  (0.02-0.02) | 0.02  (0.02-0.03) | 0.02  (0.03-0.04) | 0.02  (0.03-0.03) | 0.04  (0.05-0.06) | 0.07  (0.07-0.08) |
|  | Coefficient of influenza | 0 | 0 | 0 | 0.01 | 0.01 | 0.01 | 0.05 | 0.02 | 0.03 | 0.03 |

a: The Scottish Index of Multiple Deprivation is derived based on seven different domains, including income, employment, health, education, skills and training, geographic access to services, crime, and housing, with the 5^th^ quintile indicating the least deprived groups in Scotland.

b: The AIC for the main models.

c: The best lags (weeks) for RSV and influenza in models.

**Supplementary Table 5.** Sensitivity analyses of RSV-associated hospital admissions by all age

| **Model** | **SIMD^a^** | **AIC** | **Adjusted R square** | **Hospital admission rate (95% CI)** |
| --- | --- | --- | --- | --- |
| **Main models** | 1st | 3099.87 | 0.82 | 1.51 (1.03,1.79) |
|  | 2nd | 2985.53 | 0.82 | 1.22 (0.68,1.52) |
|  | 3rd | 2847.33 | 0.82 | 1.06 (0.72,1.26) |
|  | 4th | 2812.74 | 0.77 | 0.86 (0.55,1.02) |
|  | 5th | 2709.27 | 0.80 | 0.76 (0.43,0.9) |
| **Using negative binomial regression model** | 1st | 3127.65 |  | 1.61(1.08,1.8) |
|  | 2nd | 3017.51 |  | 1.32(0.73,1.43) |
|  | 3rd | 2877.79 |  | 1.11(0.77,1.19) |
|  | 4th | 2853 |  | 0.91(0.54,0.99) |
|  | 5th | 2729.24 |  | 0.8(0.42,0.86) |
| **Adding RV to main models** | 1st | 3105.11 | 0.82 | 1.43 (1.01,1.67) |
|  | 2nd | 3007.60 | 0.81 | 1.17 (0.62,1.46) |
|  | 3rd | 2871.13 | 0.81 | 1.01 (0.69,1.17) |
|  | 4th | 2819.15 | 0.80 | 0.8 (0.5,0.93) |
|  | 5th | 2719.16 | 0.80 | 0.73 (0.4,0.83) |
| **Adding an interaction term between influenza positive tests and 2010/11 season** | 1st | 2963.42 | 0.89 | 1.56 (1.56,1.56) |
|  | 2nd | 2868.06 | 0.88 | 1.28 (1.28,1.28) |
|  | 3rd | 2702.44 | 0.89 | 1.09 (1.09,1.09) |
|  | 4th | 2663.34 | 0.86 | 0.89 (0.89,0.89) |
|  | 5th | 2516.44 | 0.90 | 0.8 (0.8,0.8) |

a: The Scottish Index of Multiple Deprivation is derived based on seven different domains, including income, employment, health, education, skills and training, geographic access to services, crime, and housing, with the 5^th^ quintile indicating the least deprived groups in Scotland. RV: Rhinovirus
